# The influence of anatomical shape variations of wrist bones on kinematic parameter extraction in CT scans

**DOI:** 10.1101/2025.01.31.25321391

**Authors:** Maranda Haenen, Stefan Hummelink, Eline Karstanje, Ioannis Sechopoulos, Brigitte van der Heijden

## Abstract

**Introduction:** Four-Dimensional Computed Tomography (4DCT) shows promise in diagnosing wrist pathologies such as scapholunate ligament lesions (SLL). Two parameters indicative of SLL lesions in these scans, the scapholunate angle (SLA) and the capitolunate angle (CLA), might be affected by anatomical shape variations. Therefore, this study characterizes the impact of anatomical shape variations on SLA and CLA estimates from 4DCT scans.

**Methods:** 3D CT images retrieved from 4DCT scans of healthy wrists were used. Scans were automatically segmented using an Artificial Intelligence-based algorithm. A statistical shape model (SSM) was created for each bone separately. The quality of each SSM was assessed using a leave-one-out approach by calculating the root mean squared error (RMSE). Next, local coordinate systems (LCSs) were assigned to the SSMs while shape variations were introduced. The rotational deviation of the LCSs was calculated by a combined rotation around the three axes. Finally, the SLAs and CLAs were calculated for all possible bone shape combinations.

**Results:** SSMs of the carpal bones and radius were created using 106 and 99 scans, respectively, with a maximum RMSE across all SSMs of 0.65 mm. The 95^th^ percentile of the rotational deviations of the LCSs of the carpal bones and radius were below 3°. The resulting uncertainty due to anatomical variations in the calculation of the SLA and CLA was 2-3°.

**Conclusion:** Anatomical shape variations hardly influence the SLA, CLA, and LCSs of the carpal bones and radius so the methods to estimate the LCSs can be used to estimate relevant kinematic parameters in 4DCT scans.

## Introduction

Wrist injuries, like scapholunate ligament (SLL) lesions, might cause abnormal kinematics of the wrist joint, which progress to osteoarthritis if left untreated. However, due to the complexity of the wrist joint, composed of eight carpal bones, two forearm bones, and multiple extrinsic and intrinsic ligaments, an accurate diagnosis of pathology is often challenging (1–4).

Four-Dimensional Computed Tomography (4DCT) is a promising new dynamic imaging modality for evaluating wrist pathologies associated with a kinematic change. In 4DCT, multiple 3D CT scans are acquired sequentially while the wrist is in motion. This non-invasive dynamic imaging technique with high spatial and temporal resolution allows for quantifying wrist kinematics (5–7) and has been shown to have potential value in diagnosing SLL injuries (8–11).

The diagnosis of an SLL injury in 4DCT currently relies on detecting pathologic changes in the angles between the carpal bones, like the scapholunate angle (SLA) and capitolunate angle (CLA) (12, 13). To determine the SLA, CLA, and other intercarpal angles in 3D images, a unique local coordinate system (LCS) for each bone is needed. It is essential, therefore, to develop an objective and repetitive algorithm to define the LCSs in the 3D CT images so that the angle estimations are consistent and comparable across scans. However, anatomical variations might affect the definition of the corresponding LCSs and influence the derived kinematic parameters like the intercarpal angles, leading to unreliable comparative analysis across subjects. Therefore, an objective quantitative approach is needed to characterize the size of the effect of anatomic variations on the corresponding LCSs and, subsequently, on the kinematic parameters derived from the 3D images of the 4DCT scans.

A statistical shape model (SSM) is a frequently used mathematical representation of the average anatomical shape and its variability of any anatomic part of interest within a given dataset (14–17). By assigning LCSs to an SSM, modelled to the most prominent shape variations, the effect of the naturally occurring anatomical variations on the automatic LCS placement within the wrist can be investigated. The variations of the subsequently calculated angles result in an upper limit of the accuracy of the estimations of these angles. Therefore, any detectable pathology must introduce angular variations larger than this number.

Therefore, the overall aim of this study is to determine the effect of natural bone shape variations on the accuracy of SLA and CLA estimates. Thus, new methods are introduced to automatically identify LCSs within the radius, scaphoid, capitate, and lunate; a necessary first step to determine the angles. Then, the impact of anatomical shape variations on these LCSs is assessed, quantifying their subsequent effects on the SLA and CLA estimates.

## Methods

A dataset of 3D CT images retrieved from 4DCT scans of healthy wrists was used to investigate the influence of anatomical shape variations on the orientation of LCSs. For this, an SSM was created for each bone separately and then used to quantify the anatomical variations in the set. Next, LCSs were assigned to the shape models while shape variations were introduced to simulate the effect of anatomical variation on the orientation of the LCSs and the carpal angle calculations. Figure 1 shows a flowchart of the followed methodology.

**Figure 1:**
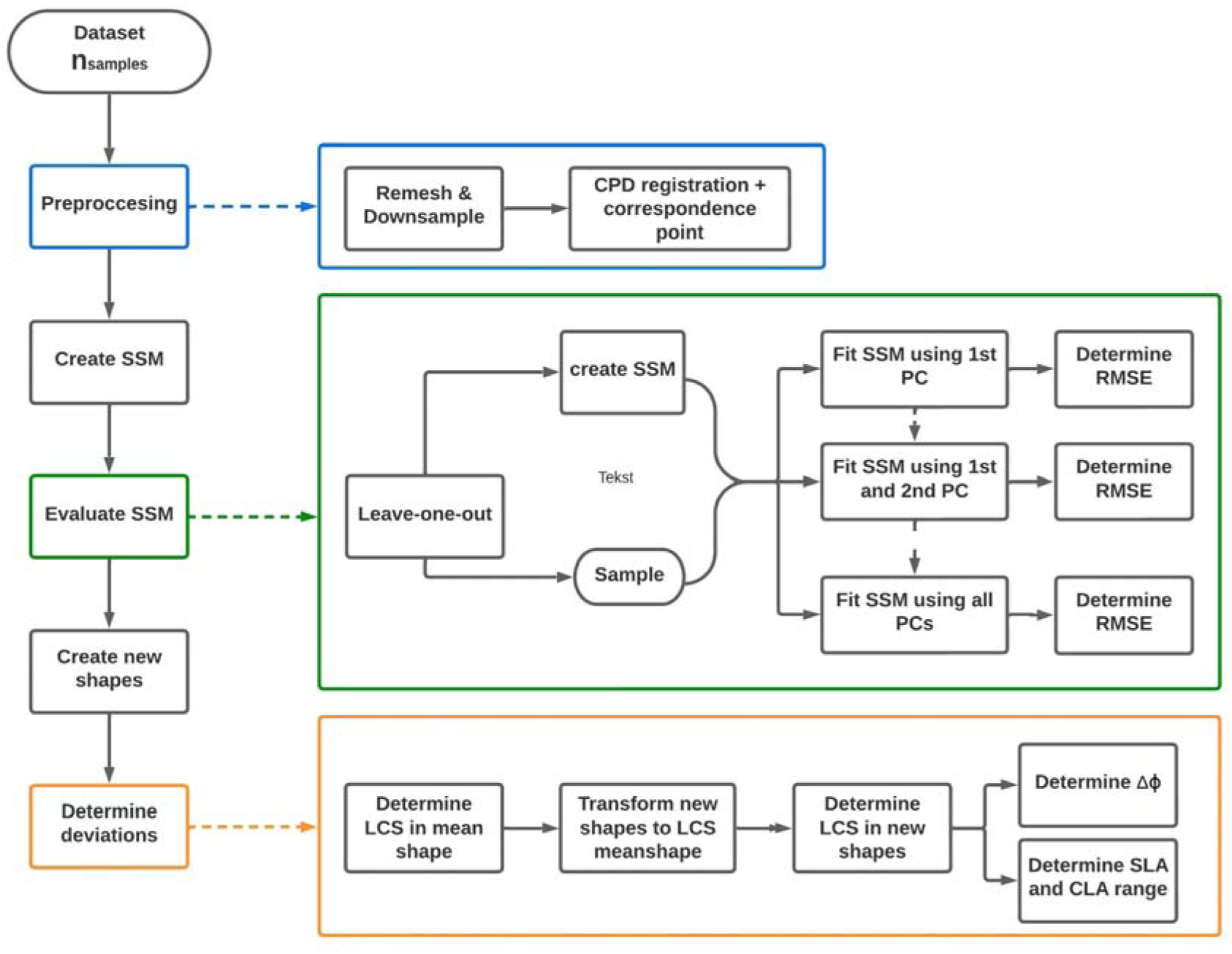
flowchart of the followed methodology. In the blue rectangle is a more detailed explanation of the preprocessing of the data, in the green one a more detailed explanation on how the quality of the SSM was assessed, and in the orange one a more detailed explanation on how the rotational error was determined.

### Dataset

The dominant hand of healthy subjects and contralateral healthy wrists of patients with no history of wrist trauma was scanned in the RadboudUMC using a clinical 320-channel CT system (Aquilion ONE Prism, Canon Medical Systems, Otawara, Japan). Ethical approval was given by the local ethical committee (ABR Number: NL72518.091.19 and NL84487.091.23). Out of the entire 4DCT scan for this study, only the initial 3D scan, during which the wrists were held static, was extracted. Scans were acquired with a helical scan, collimation 0.5 x 80 mm, pitch 0.637, rotation time 0.5 s, tube voltage 80 kV, and tube current determined automatically per patient by setting the automatic exposure control to achieve an SD of 20. Scans were reconstructed using an adaptive iterative reconstruction algorithm (AIDR 3D, Canon Medical Systems), and the average reconstructed voxel dimensions were 0.44 mm x 0.44 mm x 0.35 mm. The scans were automatically segmented using the AI algorithm created by Teule et al. and stored as surface meshes (18).

### Statistical Shape Model

The SSM was created for every bone separately using identical steps. The segmented meshes were re-meshed using anisotropic remeshing to improve mesh quality and downsampled (10000 vertices for the radius, 3000 for the carpal bones) using commercial software (MATLAB version: 9.13.0 (R2022b), The MathWorks Inc., Natick, Massachusetts). All meshes were registered and scaled to the position and overall size of their corresponding bone (i.e., radius, scaphoid, lunate, and capitate) of one randomly chosen wrist using a rigid coherent point drift (CPD) algorithm (19). Principal component analysis (PCA) was performed to identify the main variation modes, now referred to as principal components (PC) in the dataset, expressing each shape as a linear combination of these PCs. A leave-one-out approach was used to determine the generalization ability of the model. The SSM was created n_samples_ times, excluding one different sample from the set on each iteration. Then, the generalization ability was calculated by fitting the SSM to the one left-out sample and determining the root mean square error (RMSE). The fitting was repeated by progressively increasing the number of PCs included, starting with the first PC only and finally using all PCs. New shape meshes were created by setting the values of all PCs to their mean and then varying one at a time by changing the value of the PC from -2σ to +2σ in 0.2σ steps.

### Local coordinate systems and carpal angles

For the new shapes obtained from the previous step, unique LCSs were determined using different algorithms for each bone. Subsequently, the SLA and CLA were calculated. All algorithms are developed in MATLAB; a more detailed description is provided below.

#### Radius

The LCS of the radius was established using inertia tensor calculations and eigenvector analysis using the method described by de Roo et al. (20). The origin of the LCS was established between the scaphoid and lunate fossae. The long axis of the radius was defined as the z-axis of the LCS. The x-axis was set to pass through the radial styloid, which was projected onto a plane orthogonal to the z-axis, and the y-axis was defined as the cross product of the z- and x-axis, pointing in a volar direction.

#### Scaphoid

The LCS of the scaphoid was established using PCA and the direction vector of the three principal components as the direction vectors of the LCS. The first principal component passed through the proximal and distal poles of the scaphoid. The origin of the LCS was defined as the centre-of-mass (COM) of the scaphoid. To uniformly define the positive axes, the directions of the axes were corrected using the LCS determined for the radius.

#### Lunate

The LCS of the lunate was established using the two lunate cornua as landmarks. The x-axis passes from the COM of the lunate perpendicular to the line between the two cornua. A 2D plane was constructed using COM and two landmarks. The y-axis was defined as the normal vector of that plane. The z-axis was defined as the cross-product of the x and y axes. The origin of the LCS was defined as the COM of the lunate.

#### Capitate

For the LCS of the capitate, a sphere was fitted in its proximal half. The x-axis was defined as the direction from the centre of the sphere towards the COM of the capitate. For the y-axis, the normal plane of the first direction vector was defined. Subsequently, a PCA was performed on all vertices of the capitate, and the direction vector of the second principal component was projected on the normal plane of the x-axis. The z-axis was defined as the cross-product of the x- and y-axis. The origin of the LCS was defined as the COM of the capitate.

### Rotation of local coordinate systems

An LCS was determined for the mean shape of each bone, and all newly created shapes were transformed to that coordinate system. LCSs were determined for the new shapes, and the direction vectors were considered a rotation matrix from the mean shape to the new shape, making the mean shape the reference shape. This rotation matrix can be described as a rotation around the z-axis (*Δφ_y_*) followed by a rotation around the y-axis (*Δφ_z_*), and a rotation around the x-axis (*Δφ_x_*), in that order. The total rotational deviation is then given by 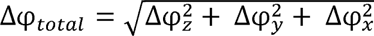.

### Carpal angle range

The SLA and CLA were calculated for all possible combinations using the new capitate, scaphoid and lunate shapes. The sagittal plane was defined as the plane constructed by the y- and z-axis of the LCS in the mean shape of the radius.

## Results

### Dataset

In total, 106 wrists from 106 subjects were scanned (52% female, 72% right wrists, age range [18y – 64y]). In seven scans, the proximal part of the radius was outside the field of view (FOV), leading to exclusion from the SSM model of the radius. This resulted in n_samples_ = 99 for the radius and n_samples_ = 106 for the scaphoid, lunate, and capitate.

### Statistical shape model

For every SSM, the total number of PCs, the number of PCs to describe 95% of the shape variances, and the minimal RMSE are shown in Table 1. The complete quality metrics of the SSMs are shown in Figure S1 in the online supplements.

**Table 1:** Quality metrics of the statistical shape models for all four bones.

| Bone | Total number of PCs | Number of PCs<br>describing 95% variance | RMSE [mm] |
| --- | --- | --- | --- |
| Radius | 98 | 74 | 0.65 |
| Scaphoid | 105 | 79 | 0.35 |
| Lunate | 105 | 82 | 0.27 |
| Capitate | 105 | 83 | 0.39 |

Abbreviations: PC, principal component; RMSE, Root Mean Square Error.

### Local coordinate systems and carpal angles

All four bones with LCS and the definition of the carpal angles are depicted in Figure 2.

**Figure 2:**
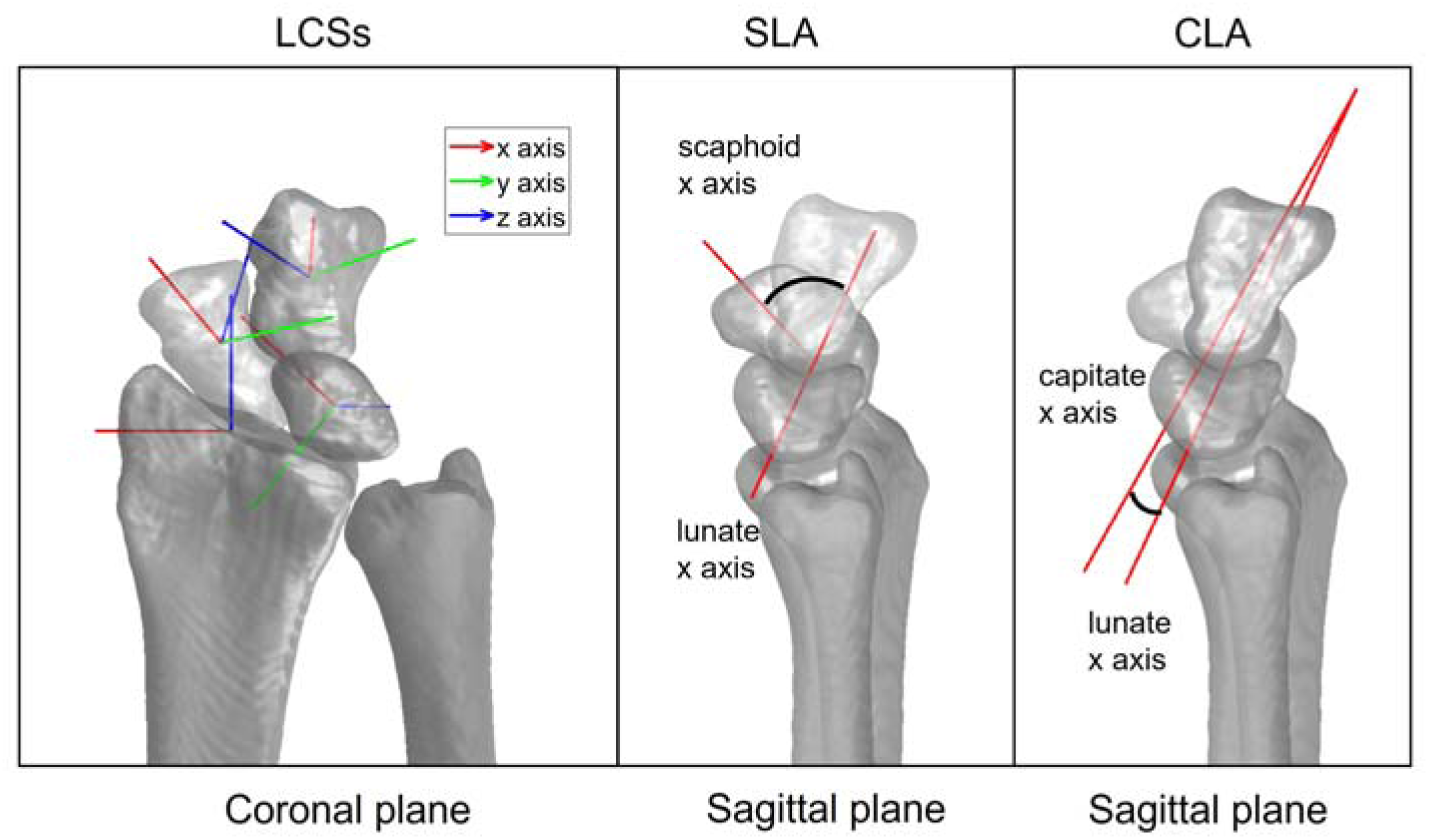
Local coordinate systems determined for the 3D images of the radius, scaphoid, lunate, and capitate depicted in an anteroposterior view/coronal plane (left) and the definition of the carpal angles SLA (middle) and CLA (right) in the sagittal plane.

### Rotation of local coordinate system

The median and 95^th^ percentile confidence interval (CI) of the for the four bones are given in Table 2. Rotational deviations are very low for the three carpal bones and the radius. The per principal component can be seen in Figure 3 and the separate rotations around each axis,, and can be seen in Figure S2, both in the online supplements. The new shapes and LCSs determined for them can also be seen in the videos S1-4 included in the online supplement.

**Table 2:**
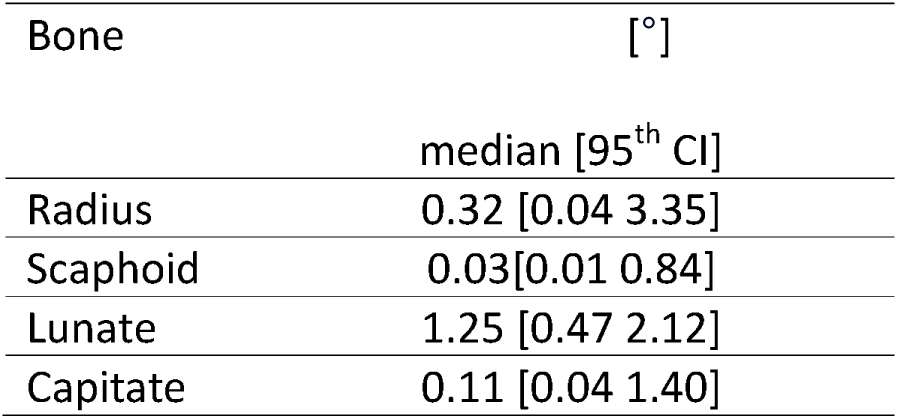
Rotational deviation of the local coordinate systems for all four bones.

| Bone | [°] |
| --- | --- |
|  | median [95<sup>th</sup> CI] |
| Radius | 0.32 [0.04 3.35] |
| Scaphoid | 0.03 [0.01 0.84] |
| Lunate | 1.25 [0.47 2.12] |
| Capitate | 0.11 [0.04 1.40] |

**Figure 3:**
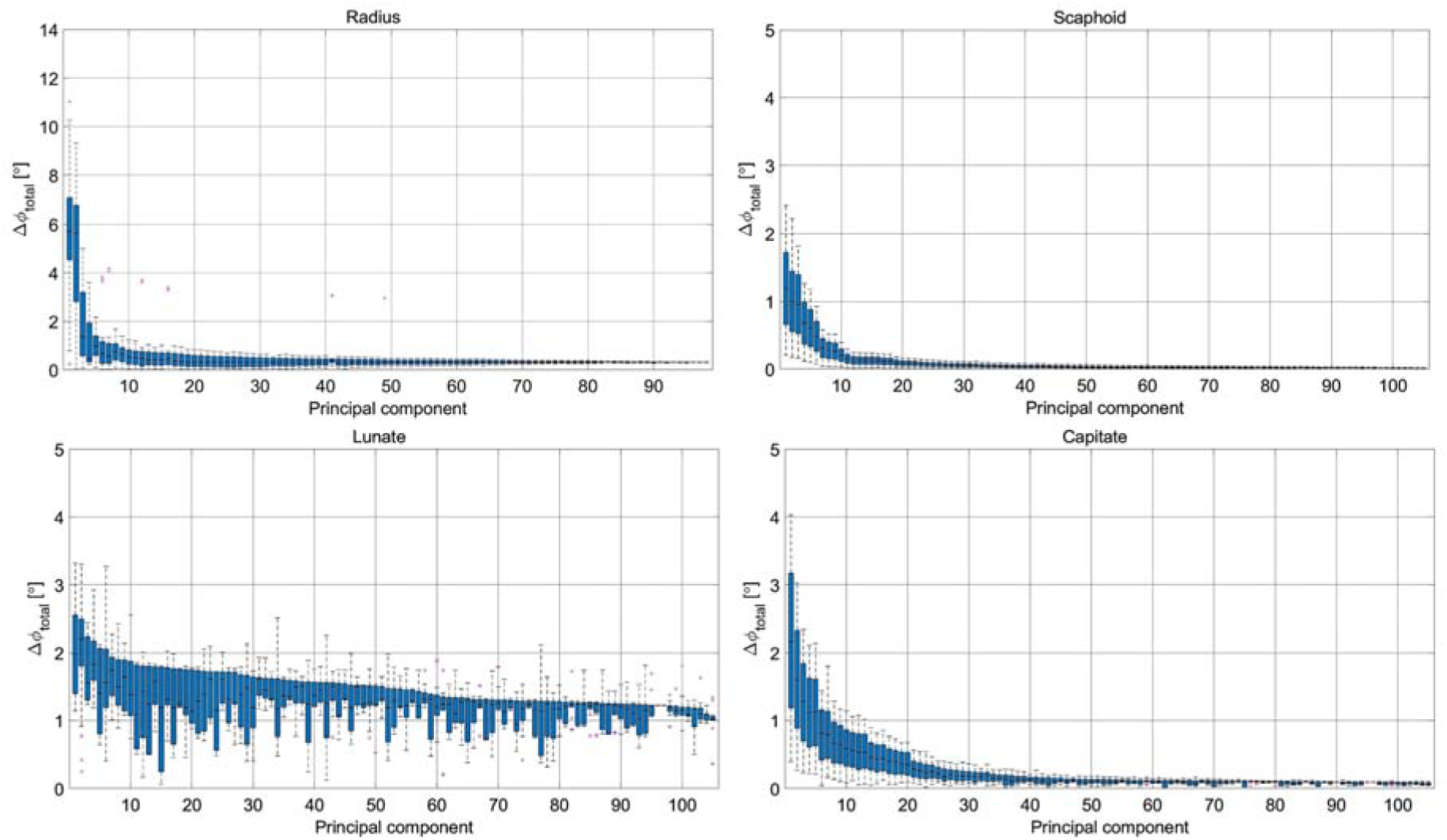
Visualization of the influence of anatomical variation on the calculated LCSs in the radius, scaphoid, lunate, and capitate. The figure shows the rotations of the LCS on the perturbed anatomy of the corresponding principal component* with respect to the coordinate system of the mean shape.** * the principal components are ordered based on the (left to right from high-to-low error) ** the limits of the y-axis are different for the graph of the radius

### Carpal angle range

For the carpal bones, 2,100 new shapes were created per bone, resulting in over four million possible scaphoid-lunate and capitate-lunate combinations. The SLA and CLA range and distributions are shown in histograms in Figure 4, together with the corresponding median and 95^th^ percentile CI. Both the SLA and CLA range are narrow, below 3°.

**Figure 4:**
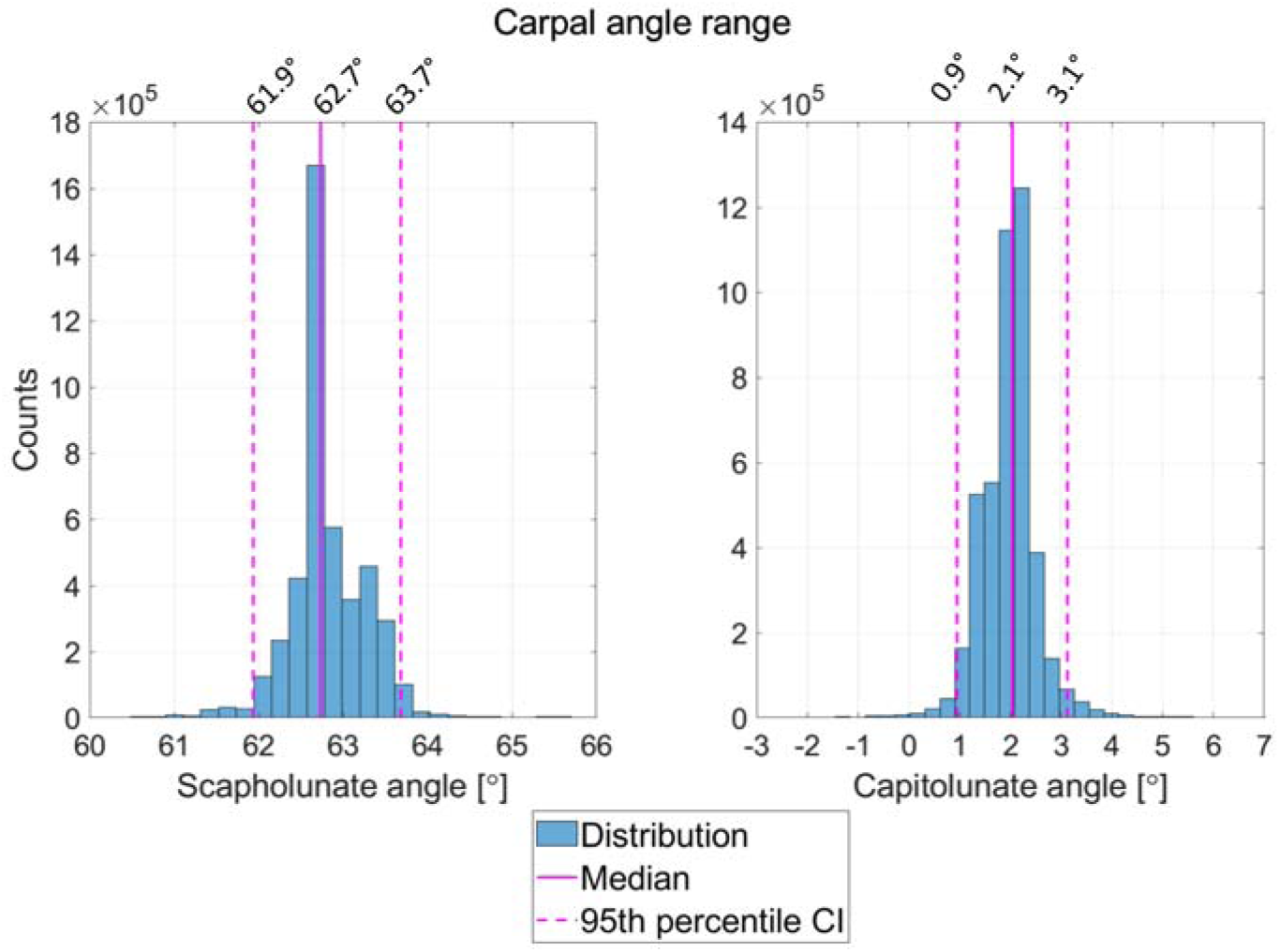
Distributions of the carpal angles calculated in over four million possible scaphoid-lunate and capitate-lunate combinations.

## Discussion

Several studies have demonstrated the potential value of 4DCT in diagnosing wrist injuries like SLL lesions. An automated assessment of kinematic parameters such as carpal angles is essential for 4DCT to be clinically feasible. For the calculation of the carpal angles in 3D images, LCSs for the carpal bone of interest must be defined. This study developed a method to automatically determine LCSs, followed by calculating carpal angles (SLA and CLA) of wrist bones. In addition, the influence of anatomical shape variations on the LCSs and the carpal angles was investigated.

Our findings show that the range of possible variation in the carpal angle estimations due to normal bone shape variability for the SLA and CLA was within 2-3°. This means that anatomical variations do not significantly influence the extraction of these kinematic parameters using the proposed LCSs. This is important in the future when comparing healthy wrists to pathological wrists. As expected, the median values for SLA and CLA are within the reference values of healthy wrists found in the literature since the radiology guidelines were considered when creating the algorithms to determine the LCSs (2). Furthermore, our findings suggest that LCSs defined in the scaphoid, lunate, and capitate are relatively insensitive to anatomical shape variations, with deviations in *Δφ_total_* below 3° (95^th^ percentile CI) and 5° overall. The radius results presented in Table 2 are similar to those of the carpal bones. However, in Figure 3, it can be seen that two out of 98 PCs result in larger deviations, with a maximum of 11°, which occur primarily around the z axis. These two PCs represent the torsion of the bone in terms of anatomical shape variations. Rotation of the LCSs in those cases was therefore expected.

All SSMs were considered of adequate quality. The literature presents comparable outcomes in evaluating the quality metrics and generalization ability of the radius, scaphoid, and lunate SSMs (14–16, 21). No comparative analysis was identified regarding the capitate, but the results obtained here align with those observed for the other two carpal bones.

One limitation of the study is the potential lack of realism of the bone shapes generated to determine the resulting variability in the LCSs and estimated SLAs and CLAs. This limitation is a result of two simplifications; that each individual bone shape was modified by varying the value of one PC at a time, and that the PC values of each bone in one wrist are independent of each other. However, this limitation probably resulted in more extreme shape variations than those that would actually be encountered, therefore testing the methodology in a worst-case scenario. Even in these extreme shapes and extreme shape combinations of the bones the SLA and CLA do not vary substantially.

The method to automatically determine LCSs developed in this study is the first step in analysing the effect of anatomic variation in bones on wrist kinematics determined in 4DCT-obtained images. For now, it was decided to capture all shape variations of the lunate in one model to keep it compact. However, numerous studies have underscored the impact of the lunate type on wrist kinematics. In the future it could be interesting to differentiate between the two types in this shape analysis (22, 23). Moreover, it might be interesting to assess the effect of anatomical shape variations on different methods of determination of LCSs, for example, the one proposed by Coburn et al. (24).

In conclusion, we developed a method to extract LCSs of wrist bones and estimated relevant kinematic parameters in 4DCT scans. This study showed that the anatomical shape variations hardly influence the SLA and CLA extraction that depend on the LCS estimations. The LCS determined in the radius is sensitive to rotations around the z axis due to the bone torsion, as expected, but is hardly influenced by other shape variations. The impact of this depends on the kinematic parameter of interest.

## Data Availability

All data produced in the present study are available upon reasonable request to the authors

## Notes

### Competing Interest Statement

The authors have declared no competing interest.

### Funding Statement

This study did not receive any funding

### Author Declarations

All subjects gave informed consent for inclusion before participating in the study. The study was conducted in accordance with the Declaration of Helsinki, and the protocol was approved by the Ethics Committee METC Oost-Nederland (ABR Number: NL72518.091.19 and NL84487.091.23).

