## Supplemental file for "The influence of anatomical shape variations of wrist bones on kinematic parameter extraction in CT scans"

**Online supplement**

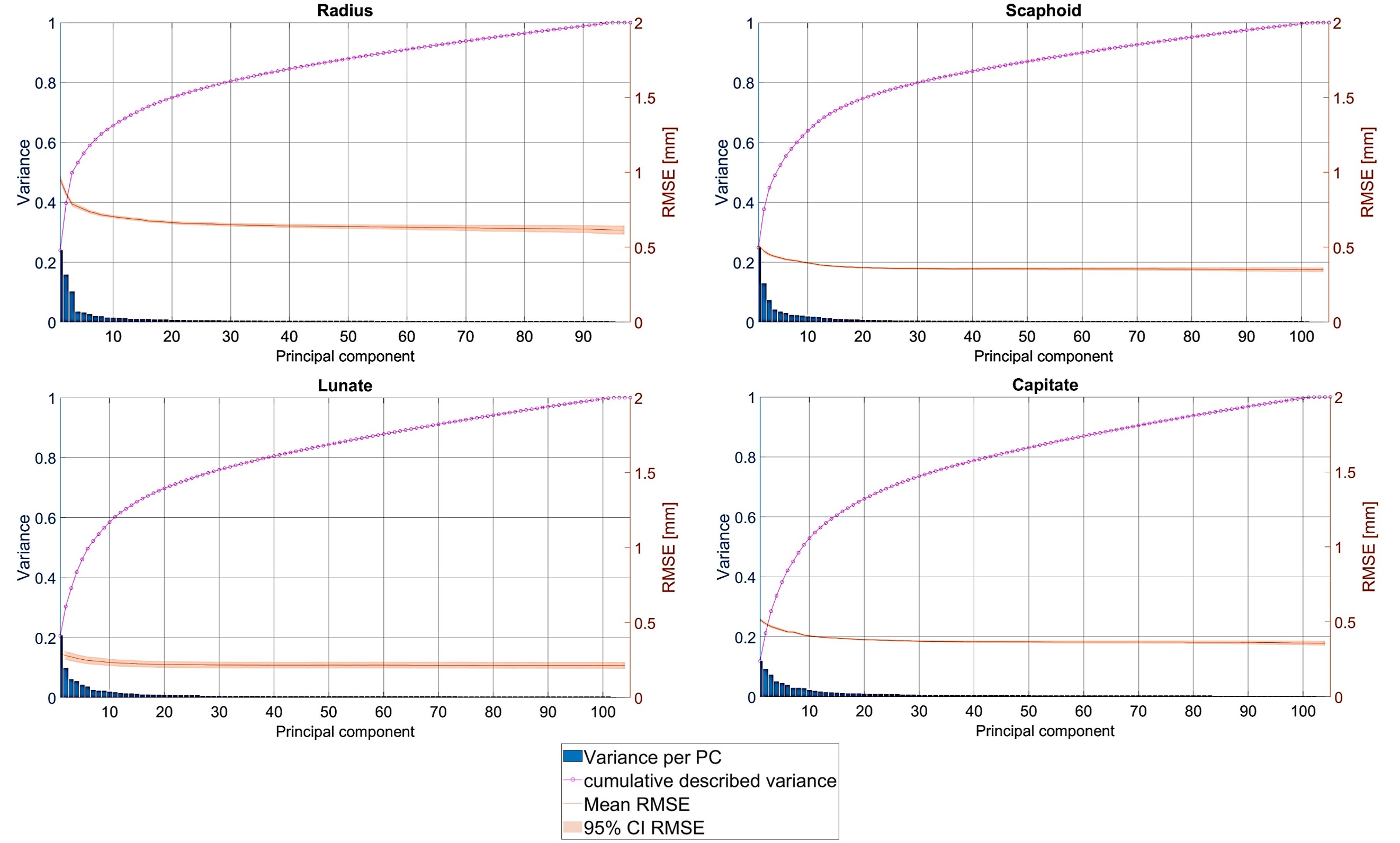

**Figure S1**: Quality metrics of the statistical shape models, as given by the cumulative described variance and the generalization ability of the radius, scaphoid, lunate, and capitate statistical shape model.

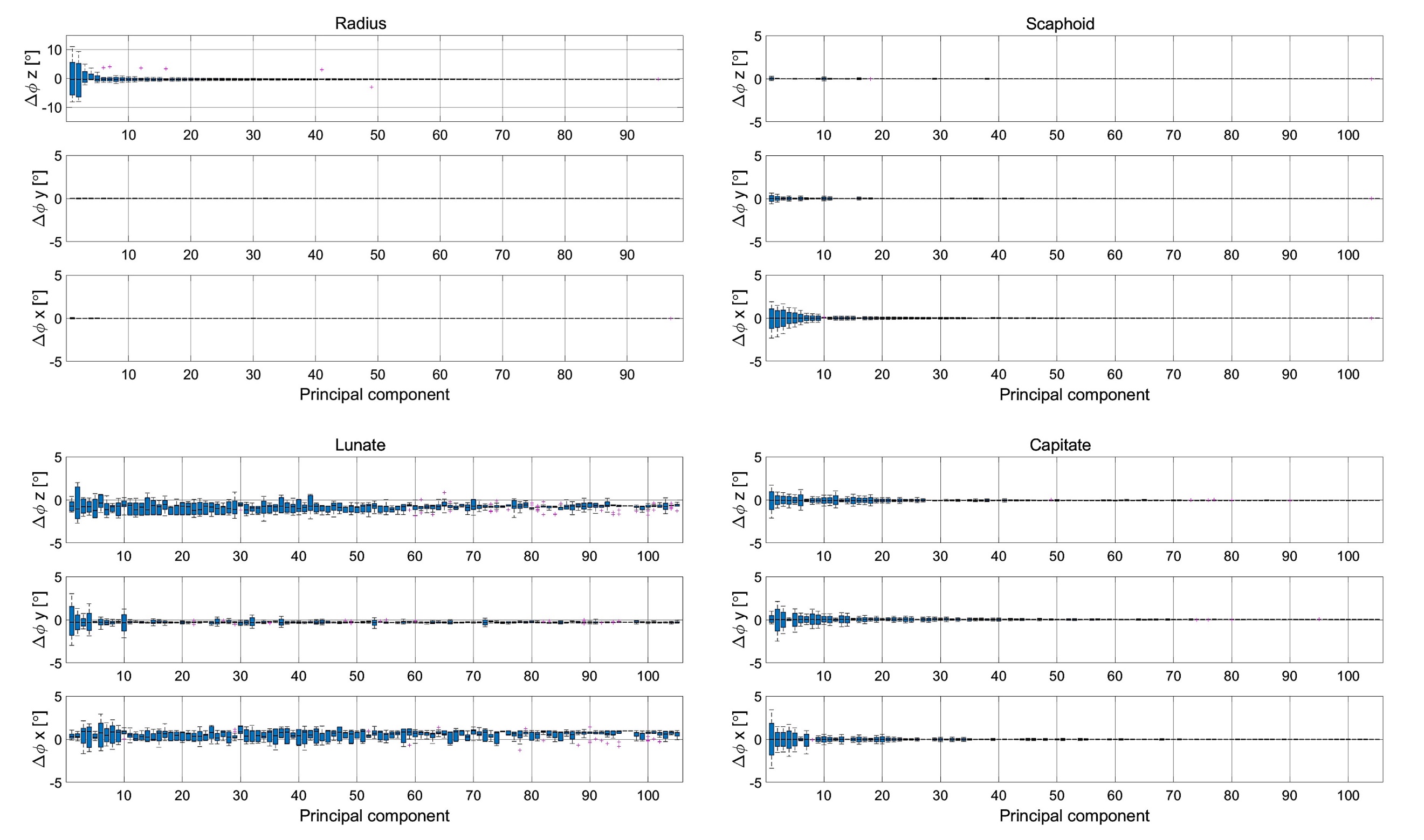

**Figure S2:** Visualization of the influence of anatomical variation on the calculated LCS in the radius, scaphoid, lunate and capitate. The figure shows the rotations of the LCS around the three axes on the perturbed anatomy of the corresponding principal component* with respect to the coordinate system of the mean shape.**

* the principal components are ordered based on the $\Delta\varphi_{total}$ (left to right from high-to-low error)

** the limits of the y-axis are different for the graph of the radius (only for rotations around the z axis)

**Video’s online supplement**

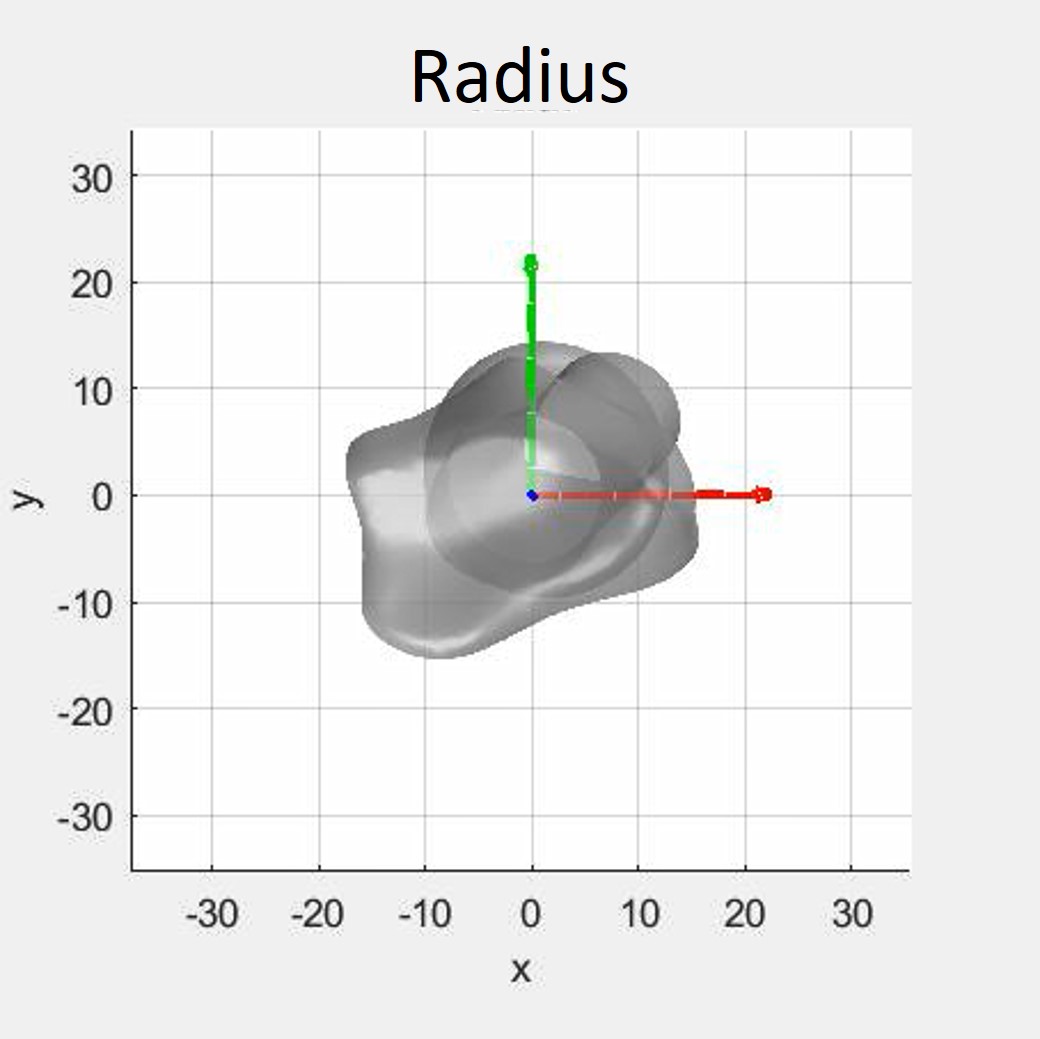

**Video S1:** Video_Radius_LCS_range

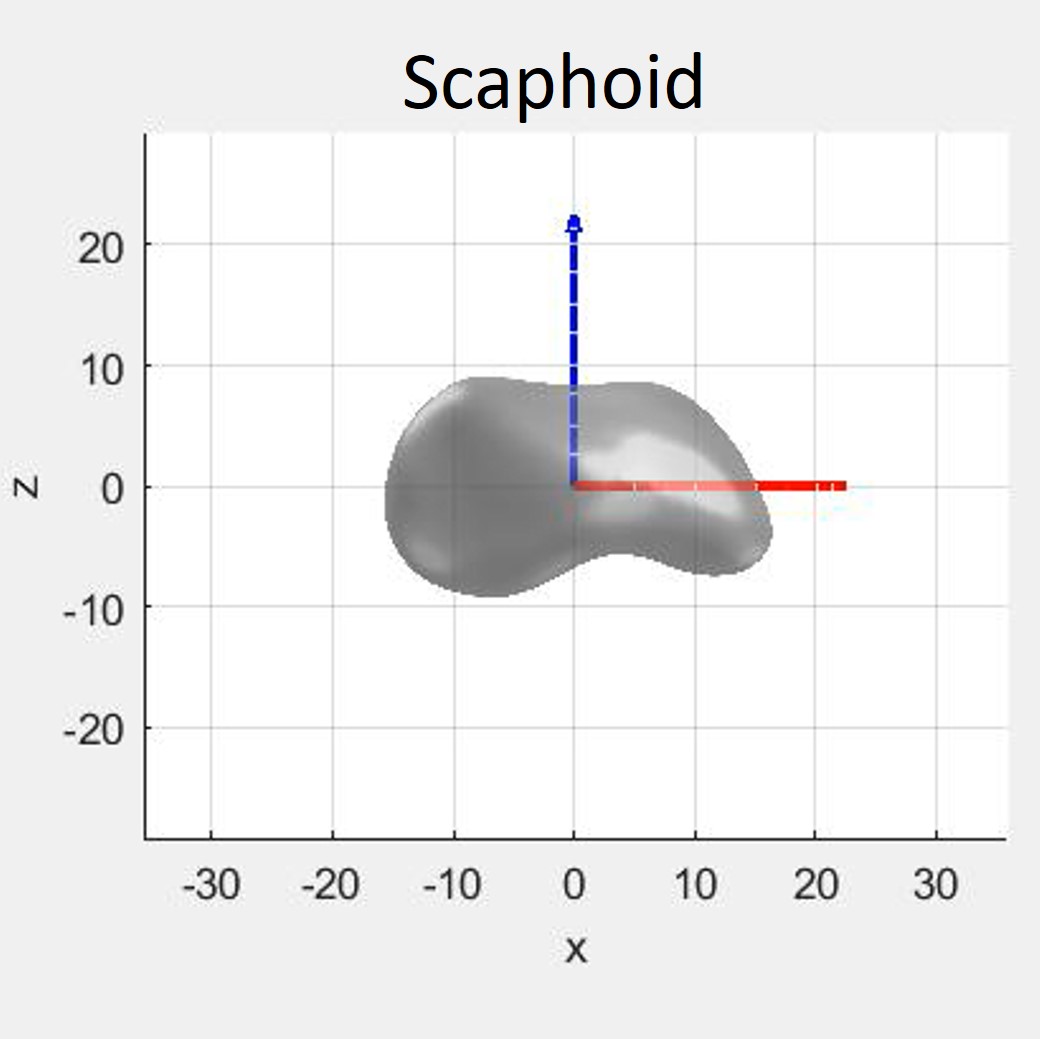

**Video S2:** Video_Scaphoid_LCS_range

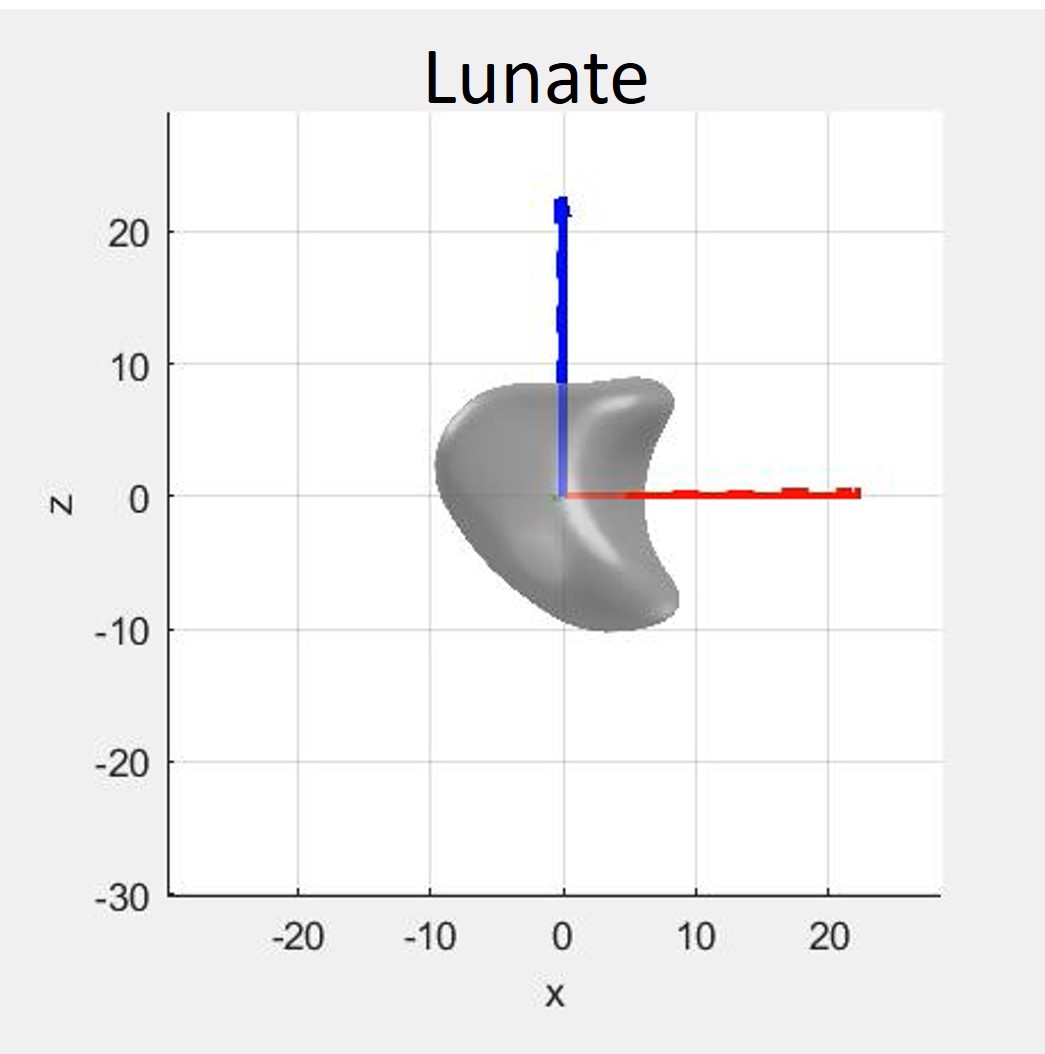

**Video S3:** Video_Lunate_LCS_range

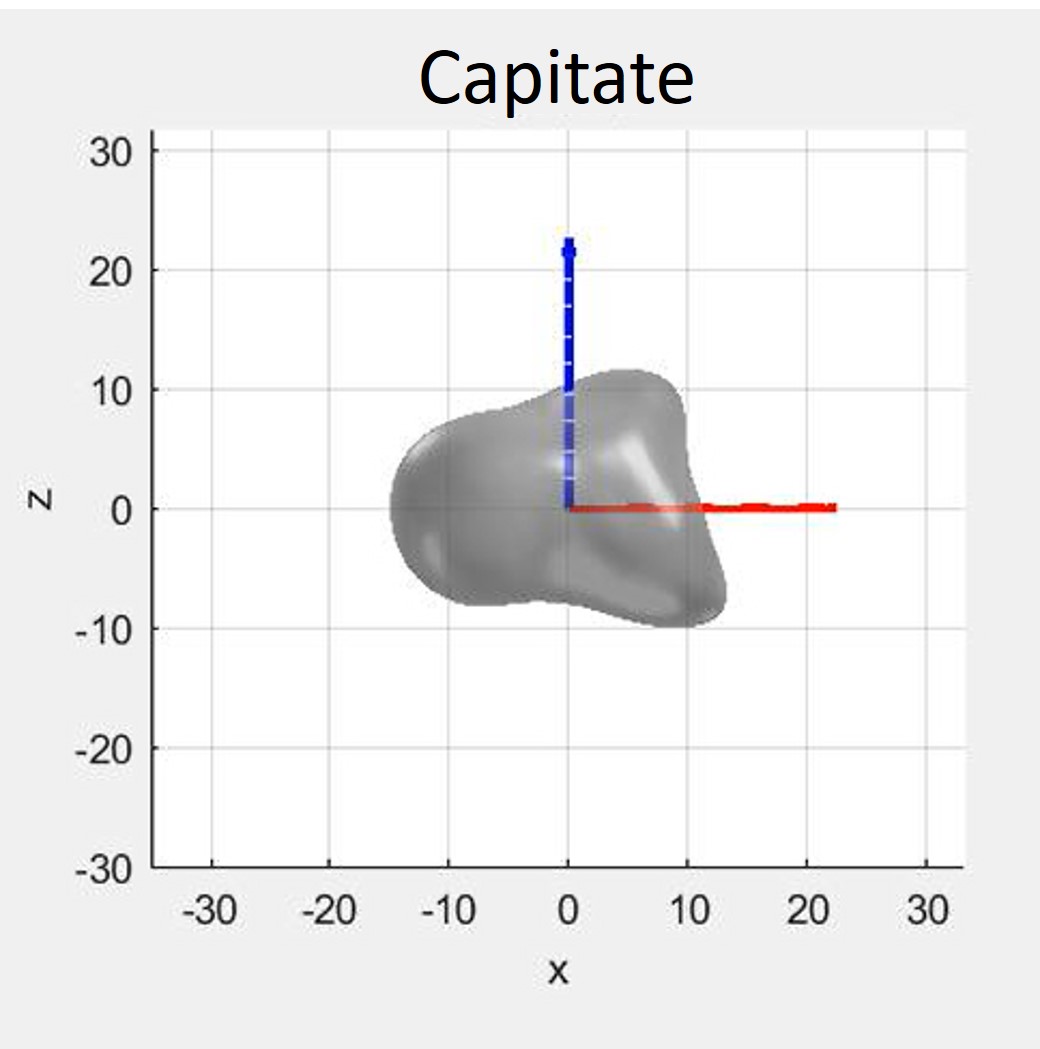

**Video S4:** Video_Capitate_LCS_range
